# Efficacy and Safety of FLOT regimen vs DCF, FOLFOX, and ECF regimens as Perioperative Chemotherapy Treatments for Resectable Gastric Cancer Patients

**DOI:** 10.1101/2021.01.26.21250550

**Authors:** Pegah Farrokhi, Alireza Sadeghi, Mehran sharifi, Payam Dadvand, Rachel Riechelmann, Azadeh Moghaddas

## Abstract

**Purpose:** This study aimed to compare the efficacy and toxicity of perioperative chemotherapy regimens including ECF, DCF, FOLFOX, and FLOT to identify the most effective chemotherapy regimen with less toxicity.

**Method:** This retrospective cohort study(2014-2021) was based on 152 eligible resectable gastric cancer patients who had received one of the perioperative chemotherapy regimens including ECF, DCF, FOLFOX, or FLOT, and followed for at least two years. The primary endpoint of this study was Overall Survival (OS), Progression-Free Survival (PFS), Overall Response Rate (ORR), and R0 resection. We also considered toxicity according to CTCAE (v.4.0) criteria as a secondary endpoint.

**Results:** Of included patients, 32(21%), 51(33.7%), 37(24.3%), and 32(21%) had received ECF, DCF, FOLFOX and FLOT, respectively. After the median 30 months follow-up, overall survival was higher with the FLOT regimen in comparison with other regimens (hazard ratio [HR] = 0. 276). The median OS of the FLOT regimen was 39 months. Besides, the median OS was 28, 25, and 21 months for DCF, FOLOFX, and ECF regimens, respectively. Moreover, a median PFS of 24, 18, 17, and 14 months was observed for FLOT, DCF, FOLFOX, and ECF regimens, respectively (Log-rank <0.001). FLOT regimen showed 84. 4% ORR, was notably higher than other groups (p-value<0. 01).

**Conclusions:** For resectable gastric cancer patients, the perioperative FLOT regimen led to a significant improvement in patients’ OS and PFS in comparison with ECF, DCF, and FOLFOX regimens. As such, the FLOT regimen could be considered the optimal option for managing resectable gastric cancer patients.

## Introduction

Gastric cancer is known as one of the most fatal cancer worldwide, being the fifth and third cause of cancer-related deaths in women and men, respectively. Eastern Asia, Eastern Europe, and South America are considered regions with a high incidence rate of gastric cancer in the world[1]. Available evidence supports the beneficial effect of perioperative chemotherapy in comparison with surgery alone on the prognosis of resectable gastric cancers[2-5]. Perioperative chemotherapy has been reported to lead to higher overall survival, progression-free survival, and curative resection rate[6,7]. According to a systematic review and meta-analysis conducted in 2015, the administration of chemotherapy before and after gastric surgery has resulted in a 42% increase in the 5-year overall survival rate in comparison with a 30% reported increase by the surgery alone[7].

The best perioperative chemotherapy regimen for the management of resectable gastric cancer is still a matter of debate. In the MAGIC trial performed on 503 patients with advanced gastric cancer in 2006, the combination of chemotherapy drugs contained epirubicin, 5-fluorouracil, and cisplatin (i.e., ECF regimen) as perioperative chemotherapy was compared to surgery alone. The results indicated that the overall survival (OS) (HR for death 0.75, 95% CI: 0.60-0.93; P=0.009; 5-year survival rate, 36% *vs*. 23%) and progression-free survival (PFS) (HR 0.66; 95% CI: 0.53 to 0.81; P<0.001) were significantly higher than control group[6]. Docetaxel together with 5-fluorouracil and cisplatin (i.e. DCF regimen) is another regimen that has been considered for perioperative chemotherapy[8]. The result of the trial indicated an improved quality of life, time to progression, and OS compared with the control group received a combination of cisplatin and 5-fluorouracil[9]. The combination of 5-fluorouracil, leucovorin, and oxaliplatin (i.e. FOLFOX regimen) has been shown as a tolerable and effective perioperative regimen for resectable gastric cancer with the median improvement of up to 22 months in PFS and 29 months in OS[10,11]. In a study conducted by *Wang et al*. The perioperative administration of modified FOLFOX6 regimen containing a higher dose of leucovorin (400mg/m^2^) and 5-fluorouracil (2.8g/m^2^) in comparison with conventional FOLFOX regimen (contains 200 mg/m2 leucovorin and vs 2.6 m^2^ 5-fluorouracil)[11,12], resulted in a 50% tumor regression rate achieved in 42.9% of patients, which was suggestive for the efficacy of FOLFOX regimen while maintaining a favorable toxicity profile[12]. According to the aforementioned studies, FOLFOX regimens seem to be effective and well-tolerated in the perioperative setting for the treatment of resectable gastric cancers. Newer combination regimens including 5-fluorouracil, docetaxel, oxaliplatin, and leucovorin (FLOT regimen) have also shown advantages over surgery alone by increasing in 3-years OS up to 58.7% in patients who received FLOT regimen in comparison with 30.9% in patients only underwent surgery[13]. The superiority of the FLOT regimen against the ECF regimen has been noted in FLOT-AIO clinical trials as well[14].

In addition to the improvement of clinical outcomes, the tolerability of the regimen with regards to toxicity is another important aspect to be considered when comparing different chemotherapy regimens. For instance, the DCF regimen has been reported to result in notable toxicity, especially in terms of neutropenia that could occur in up to 80% of patients[8]. In this context, toxicities such as grade 3-4 of neutropenia or mucositis have been reported to be more frequent in patients receiving FLOT regimen compared to those receiving ECF regimen; however, overall complications in both groups were comparable[14].

To summarize, although the available evidence has shown that the administration of perioperative chemotherapy could lead to an improvement in the prognosis of the patients suffering from resectable gastric cancers[15], however, studies are needed to compare these regimens in terms of their beneficiary effects and toxicity, enabling clinicians to choose the optimal regimen. To our knowledge, there is no available head-to-head clinical trial, comparing the aforementioned common chemotherapy regimens for resectable gastric cancer patients. In this study, we aimed to evaluate and compare the efficacy and toxicity of common regimens used as perioperative chemotherapy including ECF, DCF, FOLFOX, and FLOT in resectable gastric cancers to identify the most effective chemotherapy regimen with less toxicity.

## Methods and Materials

This retrospective cohort study, conducted in Iran, was based on the follow-up of patients with resectable gastric cancers who had undergone gastrectomy and received peri-operational chemotherapy. Iran has one of the highest incidences of resectable gastric cancer in the world with an incidence rate of 15 and 8 per 100000 cases in men and women, respectively[16-18]. The study was approved by the Ethics Committee of Isfahan University of Medical Sciences with the Iranian approval ID of IR.MUI.RESEARCH.REC.1398.167 and all enrolled patients signed the consent form at the beginning of data gathering.

### Patients

We recruited all the eligible patients with resectable gastric cancer who had information in the archives of the hospital admitted to the Omid Hospital, Isfahan, Iran, between July 2016 and July 2019. Omid Hospital is a tertiary referral hospital allocated exclusively to oncology patients and affiliated to the Isfahan University of Medical Sciences. The inclusion criteria were: (i) age between 18-75 years old at the time of diagnosis, (ii) pathologic diagnosis of the resectable gastric tumor without metastasis, (iii) having undergone gastrectomy (partial or total), (iv) having received any chemotherapy regimens including ECF, FOLFOX, DCF or FLOT as the first-line chemotherapy in preoperative and postoperative chemotherapy plan, (v) being without any concurrent active malignancy. In the Omid hospital, the patient is considered for surgery when they have no or limited metastasis, with the latter defined as having “abdominal or retroperitoneal lymph node metastases only (e.g. para-aortic, intra-aortic-canal, peripancreatic, or mesenteric lymph nodes) or one incurable organ site including bilateral or unilateral Krukenberg tumors, unilateral or bilateral adrenal gland, extra-abdominal lymph node metastases, such as supraclavicular lymph node involvement, with or without retroperitoneal lymph node metastases and no clinically visible (on CT scans or ascites) or symptomatic carcinomatosis of peritoneum or pleura and no diffuse peritoneal carcinomatosis on diagnostic laparoscopy or fewer than five liver metastases, if the single organ site is the liver, with no clinically visible (on CT scans or because of ascites) or symptomatic carcinomatosis of peritoneum or pleura and no diffuse peritoneal carcinomatosis on diagnostic laparoscopy”[19].

### Chemotherapy regimens

The following perioperative chemotherapy regimens were administrated for our included patients:

1. *<u>DCF regimen</u>* containing Docetaxel (75 mg/m^2^, day 1), Cisplatin: (75 mg/m^2^, days 1), and Fluorouracil: (750 mg/m^2^/day, days 1-5).
2. *<u>ECF regimen</u>* including Epirubicin (50 mg/m^2^, day 1), Cisplatin (60 mg /m^2^, day1), and Fluouracil (200 mg/m^2^/day, continuous infusion during days 1-21).
3. *<u>FOLFOX regimen</u>* containing Oxaliplatin (85 mg/m^2^, day 1), Leucoverin (200 mg/m^2^, day 1), and Fluouracil (2600 mg/m^2^ continuous infusion over 24 hours, day 1).
4. *<u>FLOT regimen</u>* including Docetaxel (50 mg/m^2^, day 1), Oxaliplatin (85 mg/m^2^, day1), Leucoverin (200 mg/m^2^, day 1), and Fluouracil (2600 mg/m^2^ continuous infusion over 24 hours day 1).

Before and during each cycle of chemotherapy the bone marrow function was checked and all patients received filgrastim (7.5 mg/kg), 48-72 hours after the completion of chemotherapy. In the case of any grade 3-4 toxicity, the chemotherapeutic dosage of the next cycle was reduced by 20%.

### Outcome evaluation

During the follow-up period, the evaluation of the tumors’ response was carried out through the review of histopathologic reports and computed tomography (CT) scans according to the RECIST CRITERIA v 1.1[20]. The physicians performed a response evaluation every 8-12 weeks after initiation of perioperative chemotherapy during the entire follow-up period. According to these criteria, clinical response was classified based on a decrease in tumor size as following: “(i) *complete response (CR)* defined as the disappearance of tumor lesion for at least four weeks; (ii) *Partial response (PR)* defined as more than 30% decrease of the longest diameter of the tumor lesion that was lasting for at least four weeks; (iii) *Progressive disease (PD)* was defined as a 20% increase in the longest diameter of a target tumor lesion or appearance of a new lesion on imaging findings; and (iv) *stable disease (SD)* was considered for a patient who had not met the criteria of progressive disease or partial response”[20]. We defined the *overall response rate* (ORR) as the sum of CR and PR. Furthermore, the sum of CR, PR, and SD was considered as the *disease control rate* (DCR).

### Adverse events

The adverse effects of chemotherapy were evaluated according to the Common Terminology Criteria for Adverse Events (CTCAE) 5.0 criteria[21]. We focused on all reported 3-4 grades of hematological including anemia, thrombocytopenia, and neutropenia. Besides, grade 3-4 of gastrointestinal toxicities, as well as nausea, vomiting, diarrhea, and mucositis were evaluated according to CTCAE criteria.

### Endpoints

The primary endpoints of this study were Overall response rate, Overall Survival (OS), and Progression-Free Survival (PFS) associated with each chemotherapy regimen in patients who were undergone perioperative chemotherapy with resective surgery. The OS was defined as the length of time from either the date of diagnosis or the start of treatment until patients ‘death for any reason. Progression-free survival was defined as the length of time during and after the treatment until disease progression. R0 resection which indicated no microscopic remained cancer cells in primary tumor site and major toxicities (mainly grade 3–4 hematological and non-hematological adverse effects) were also considered as secondary endpoints.

### Statistical analysis

We reported and compared the continuous variables (mean ± standard deviation (SD)) and categorical variables using a one-way ANOVA test and Chi-square test, respectively. Regarding analyses of the primary outcome, we estimated OS and PFS by the Kaplan-Meier method and compared survival using the log-rank test. All aforementioned tests were considered statistically significant if they had a two-sided *p-value* less than 0.05. To calculate the crude and adjusted hazard ratio (HR) with 95% confidence interval (CI) with ECF regimen as the referent, the univariate and multivariate cox-regression analyses were applied. The adjusted HR was based on confounders such as an ECOG performance status, family history of cancer, disease histopathology, cancer stage, tumor size, having other organ metastasis, site of metastasis, resection rate, and the response to chemotherapy (defined as a progressive disease or stable disease) as a result of plausibility and associations with survival. To estimate the clinical and pathological patients’ characteristics on prognosis, first, we performed univariate logistic regression analysis for every single variable, then the covariate with p-value <0.1 were considered in multivariate logistic analyses. The results of multivariate analyses on covariates which were shown p-value <0.05 defined as statistically significant and considered as independent prognostic factors in survival. The IBM SPSS software (Chicago, IL, USA), version 25.0 was used to calculate statistical analysis.

## Results

The characteristics of patients have been shown in Table 1. Among 152 included patients, 32(24.2%), 51(33.5%), 37(24.3%), 32(21%) had received ECF, DCF, FOLFOX, and FLOT as the perioperative chemotherapy regimens, respectively. The majority of the included patients were male and the mean age of enrolled patients was 55.95 ± 12.01 years old. The mean age of the patients was significantly higher in those who received the FOLFOX regimen in comparison with the other chemotherapy regimens. Eighty-nine (58.9%) patients had a normal BMI (18-24 kg/m^2^). Eastern Cooperative Oncology Group (ECOG) performance status was less than 2 in 92.7% of enrolled patients. Eighty (52.6%) and 75(49.3%) patients had a history of smoking and Helicobacter pylori (H. Pylori) infection, successively. Most of the patients with a positive history of smoking had been received a DCF regimen in comparison with the other regimens. The tumor size of less than 4 cm was found in 64 (42.4%) patients. One hundred and thirty (85.5 %) patients had no metastatic lesion and the proximal tumor site was seen in 86 (56.6%) patients. According to the TNM classification, stage 3 (including 3a,3b) was reported in 40 (26.3%) patients and the N1 and T3 stage was recorded in 69 (45.4%) and 60 (39.4%) patients, respectively.

**Table 1.**
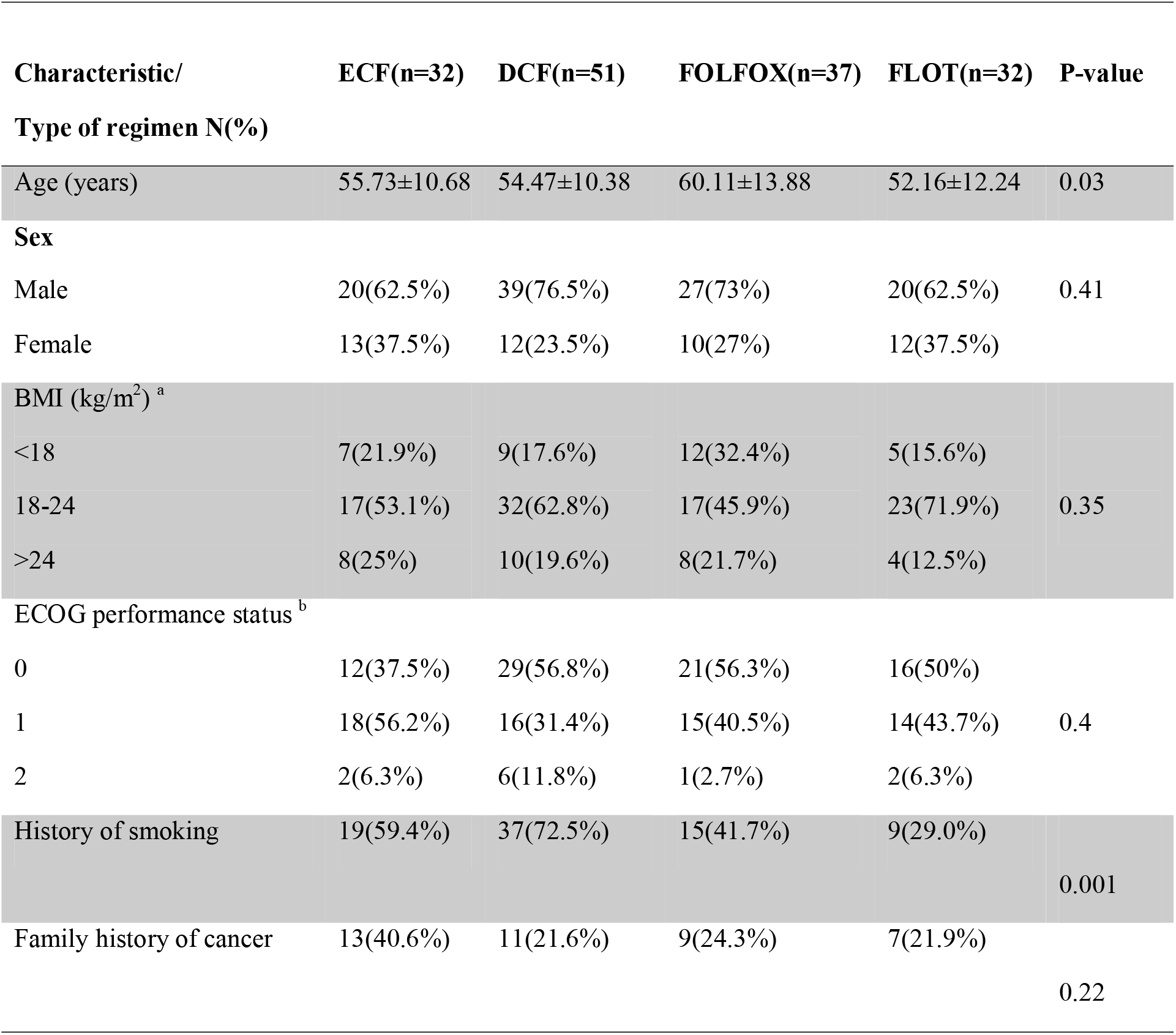
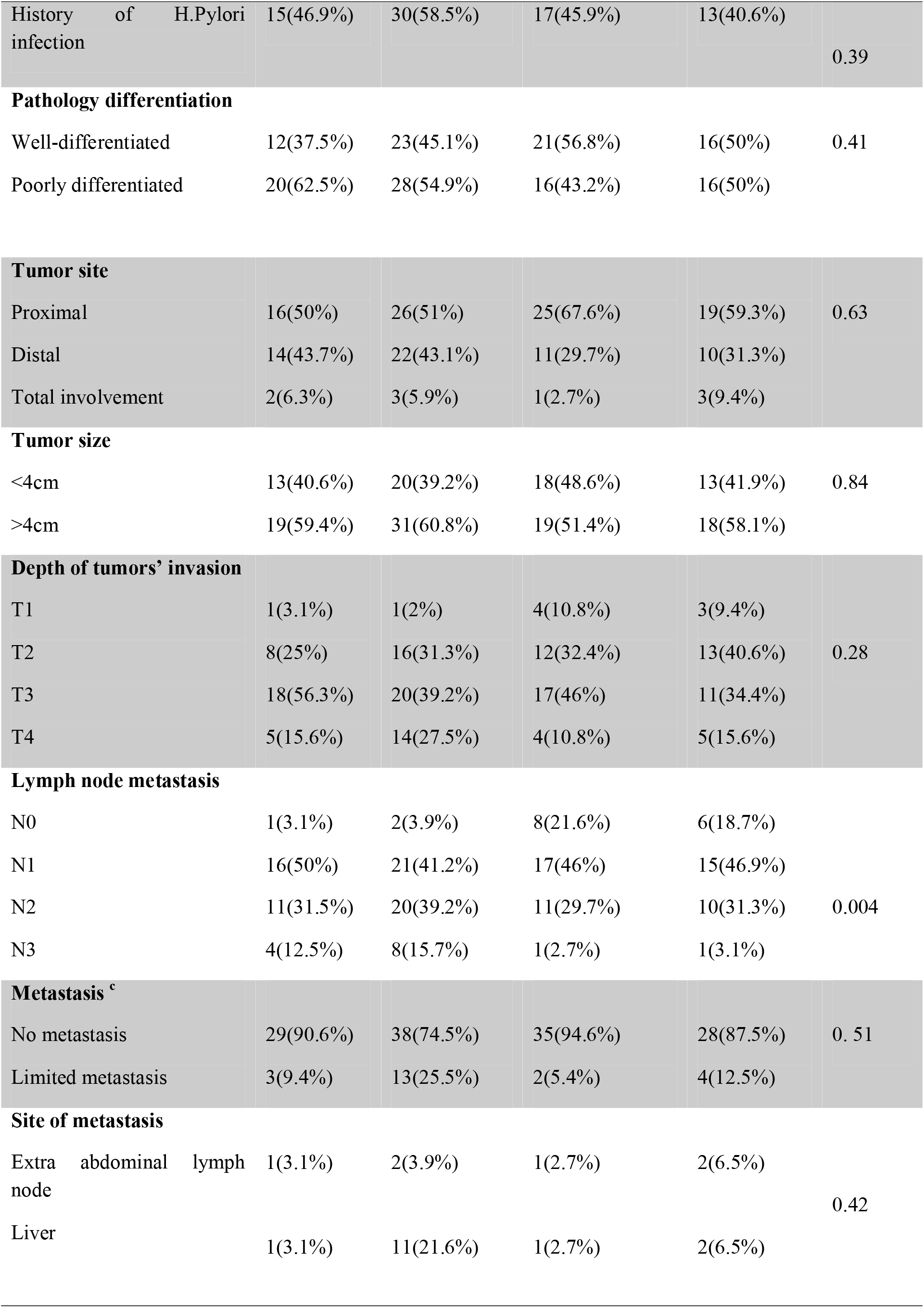
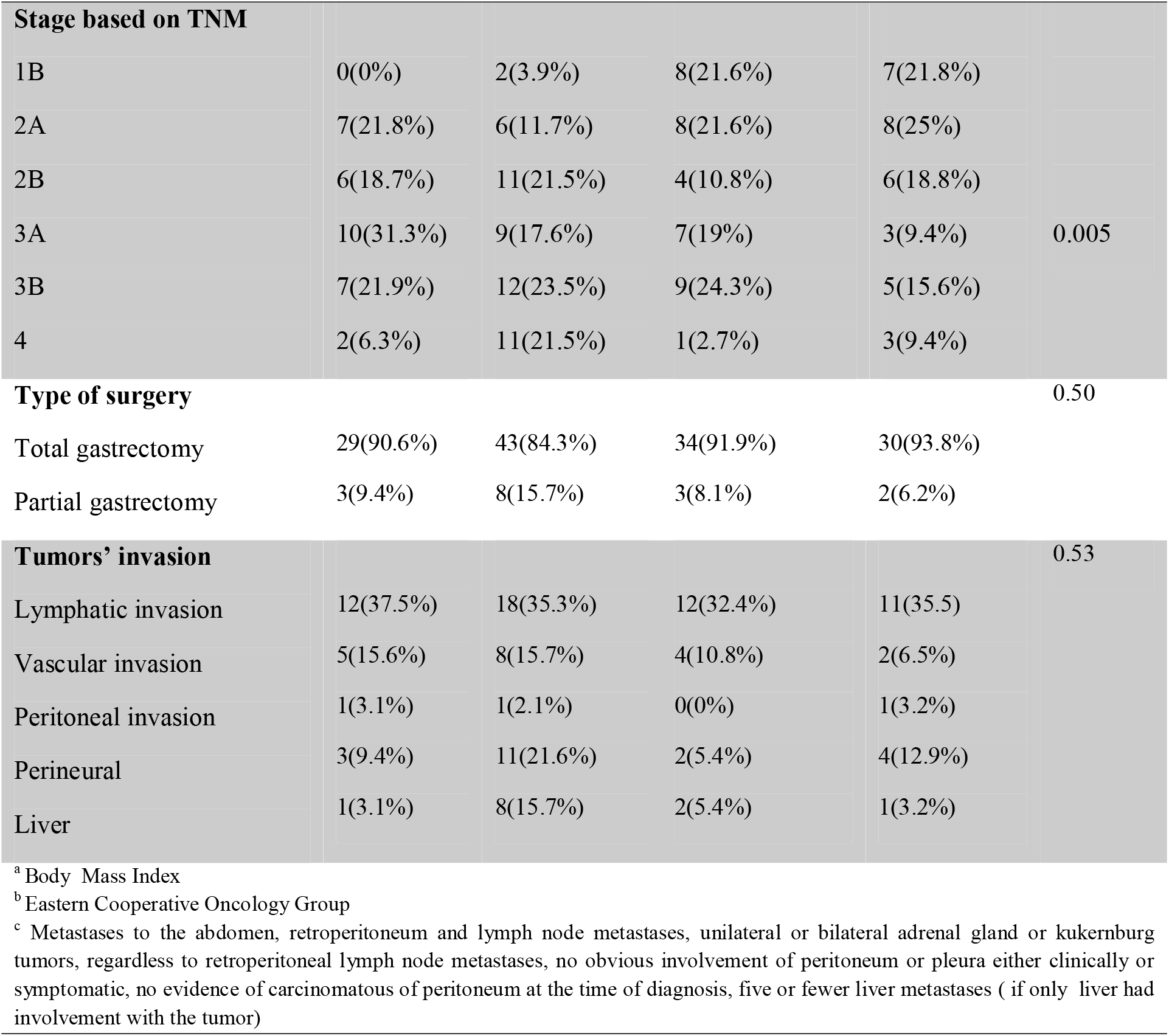
Demographic characteristics of patients.

One hundred and thirty-six (89.5%) patients underwent total gastrectomy. The most common site of invasion was lymph nodes. Free margin of surgery (R0 resection) was reached in 132 (86.6%) patients, although 20 (13.2%) of patients had a positive surgical margin either in distal or proximal tumors’ site.

### Chemotherapy efficacy

The results of response rate based on the received chemotherapy regimens were summarized in Table 2.

**Table 2.** The results of response to chemotherapy (Patients (N= 152)).

| Response/ |  |  |  |  |  |
| --- | --- | --- | --- | --- | --- |
| Type of regimen | ECF(n=32) | DCF(n=51) | FOLFOX(n=37) | FLOT(n=32) | P-value |
| N(%) |  |  |  |  |  |
| **Responserate** |  |  |  |  |  |
| PR<sup>a</sup> | 18(56.2%) | 36 (70.6%) | 27(73.0%) | 26(81.3%) |  |
| CR<sup>b</sup> | 0(0%)10(31.3%) | 3(5.9%) | 1(2.7%) | 1(3.1%) |  |
| SD<sup>c</sup> | 4(12.5%) | 7(13.7%) | 6(16.2%) | 4(12.5%) | 0.002 |
| PR<sup>d</sup> |  | 5(9.8%) | 3(8.1%) | 1(3.1%) |  |
| **Overall response rate<sup>e</sup>** | 18(56.3%) | 31(76.47%) | 28(75.7%) | 27(84.37%) | <0.001 |
| **Disease control rate<sup>f</sup>** | 28(87.6%) | 46(90.1%) | 33(89.1%) | 31(96.8%) | <0.001 |
| **Resection Rate** |  |  |  |  |  |
| **R0<sup>g</sup>** | 24(75.0%) | 48(94.1%) | 30(81.8%) | 30(93.8%) |  |
| **R1<sup>h</sup>** | 8(25.0%) | 3(5.9%) | 7(18.9%) | 2(6.2%) | 0.03 |
| **Hospitalization rate** | 12(37.5%) | 26(45.6%) | 6(16.2%) | 13(37.25) | 0.05 |
| <sup>a</sup> Partial response, <sup>b</sup> Complete response, <sup>c</sup> Stable disease, <sup>d</sup> Progression disease, <sup>e</sup> Partial response + complete response<br><sup>f</sup> Partial response + complete response + stable disease<br><sup>g</sup> No microscopic cancer cell residual in the primary tumor site<br><sup>h</sup> Microscopic cancer cell residual in the primary tumor site |  |  |  |  |  |

**Table 3.**
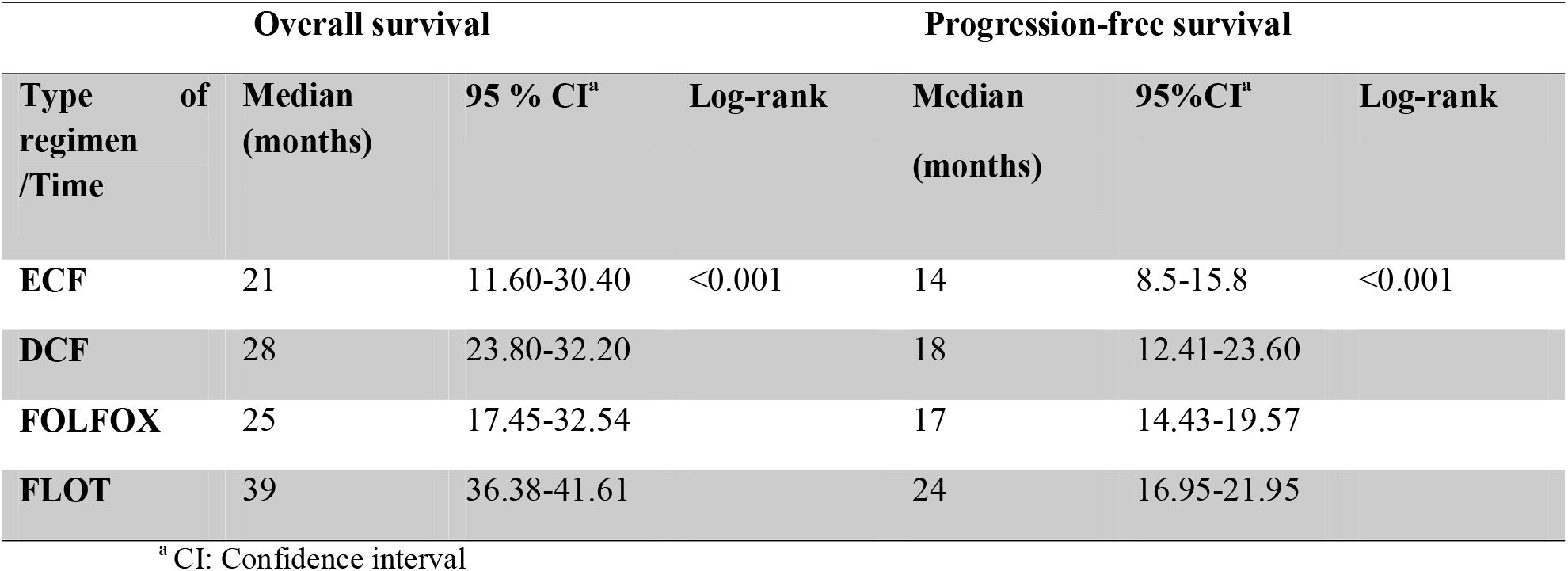
Kaplan-Meier survival analysis results.

Complete response was reported in only one patient who received FOLFOX and FLOT regimes and two patients with DCF regimens. Besides, DCR and ORR were also remarkably higher in the FLOT regimen in comparison with the other regimens (p-value<0.05). R0 resection rate was significantly higher in the FLOT regimen (93.8%) and DCF regimen (94.11%) than the other regimens (p-value=0.01). (See Table 2)

Kaplan-Meier survival analysis demonstrated that patients who received the FLOT regimen had a higher median PFS time (24 months, CI 95%=16.95-21.95, log-rank=0.001). In addition, in terms of OS time, the FLOT regimen demonstrates superiority over other regimens by 39 months overall survival times.

Regarding the comparison between other regimens, the median OS of the DCF regimen was higher than ECF and FOLFOX regimen; off note, the difference was not statistically significant (p-value=0.41). There was no significant difference between median PFS of all FOLFOX, ECF, and DCF regimens. The one-year and two-year survival rate of the FLOT regimen was calculated substantially higher in comparison with the other regimens and estimated to be 64%-28%,94%-57%, 86%-48%, 100%,97%, and 100%-96.9% in ECF, DCF, FOLFOX, and FLOT regimens, respectively.

Moreover, the results of one-year and two-years’ PFS analysis were shown to be 39%,11% for ECF regimen, 57%,34% for DCF regimen, 62%,18% for FOLFOX regimen and 91%,50% for FLOT regimen.

By considering ECF regimen as a reference regimen in cox-proportional analysis, FLOT regimen demonstrated its significant superiority in overall response (HR=0.276, p-value=0.001) and PFS (HR=0.388, p-value= 0.01) in comparison with the other regimens. Besides, subgroup analysis revealed that the DCF regimen had a significant survival rate in comparison with the ECF regimen in the crude model analysis (HR=0.755). However, when these differences were adjusted by confounders such an ECOG performance status, family history of cancer, disease histopathology, cancer stage, tumor size, having other organs’ metastasis, site of metastasis, resection rate, and the response to chemotherapy (defined as progressive disease or stable disease), the differences were not remarkable either for survival rate or for PFS. (See Table 4, 5 and Figure 1,2)

**Table 4.** Cox-regression survival analysis.

| Chemotherapy regimen | Overall Survival |  |  | Progression-Free Survival |  |  |
| --- | --- | --- | --- | --- | --- | --- |
|  | Adjusted model <sup>a</sup> |  |  | Adjusted model |  |  |
|  | HR<sup>b</sup> | 95% CI<sup>c</sup> | P-value | HR<sup>b</sup> | 95%CI<sup>c</sup> | P-value |
| ECF | 1 | - | 0.017 | 1 | - | **0.06** |
| DCF | 0.775 | 0.48-1.24 | 0.291 | 0.798 | 0.47-1.39 | **0.363** |
| FOLFOX | 0.677 | 0.39-1.15 | 0.151 | 0.810 | 0.588-1.812 | **0.449** |
| FLOT | 0.276 | 0.15-0.52 | 0.001 | 0.388 | 0.22-0.68 | **0.01** |

<sup>a</sup>Based on ECOG performance status, family history of cancer, disease histopathology, cancer stage, tumor size, and stage, having other organs' metastasis, site of metastasis, resection rate, and response to chemotherapy
<sup>b</sup> Hazard ratio
<sup>c</sup> Confidence interval

**Table 5.**
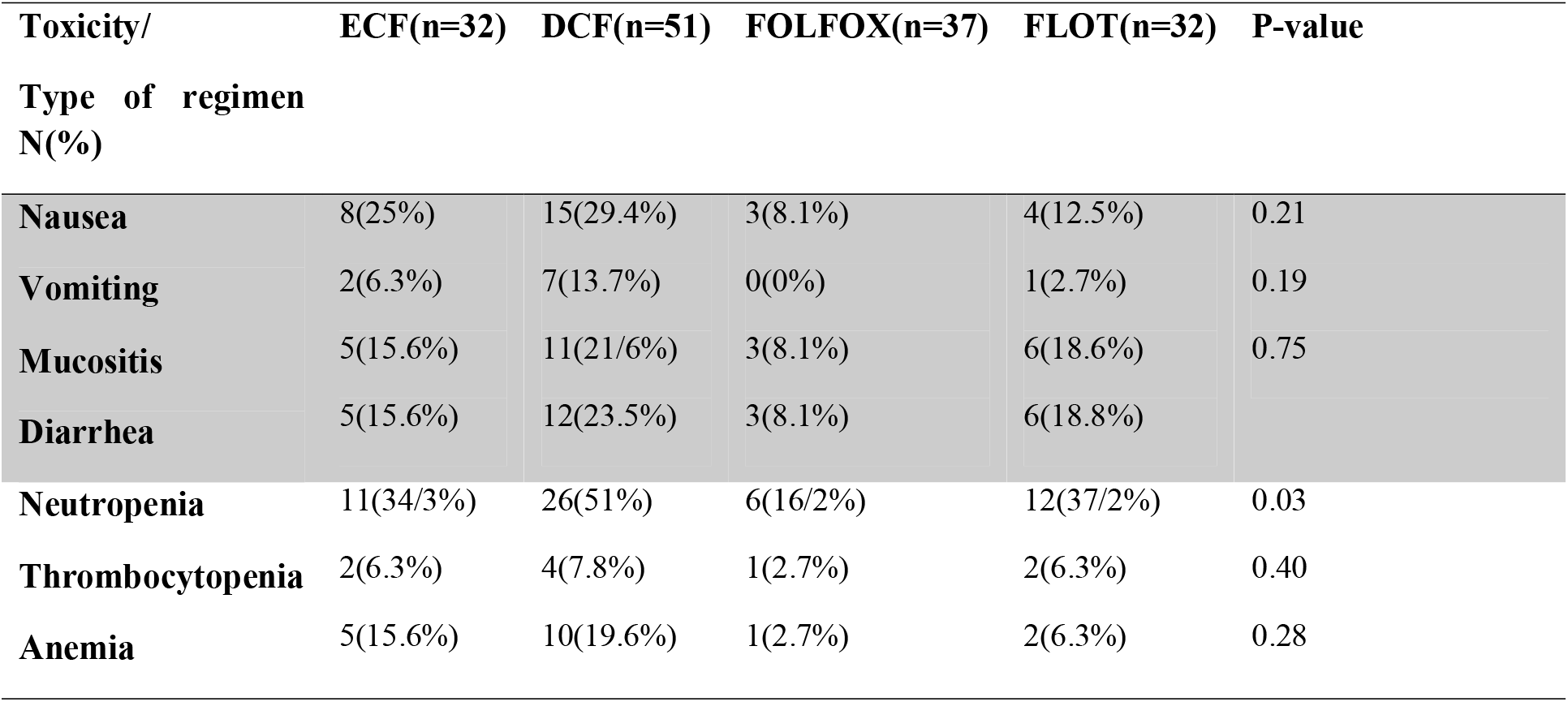
Toxic effects of chemotherapy regimens based on CTCAE criteria.

| Toxicity/<br>Type of regimen<br>N(%) | ECF(n=32) | DCF(n=51) | FOLFOX(n=37) | FLOT(n=32) | P-value |
| --- | --- | --- | --- | --- | --- |
| **Nausea** | 8(25%) | 15(29.4%) | 3(8.1%) | 4(12.5%) | 0.21 |
| **Vomiting** | 2(6.3%) | 7(13.7%) | 0(0%) | 1(2.7%) | 0.19 |
| **Mucositis** | 5(15.6%) | 11(21.6%) | 3(8.1%) | 6(18.6%) | 0.75 |
| **Diarrhea** | 5(15.6%) | 12(23.5%) | 3(8.1%) | 6(18.8%) |  |
| **Neutropenia** | 11(34.3%) | 26(51%) | 6(16.2%) | 12(37.2%) | 0.03 |
| **Thrombocytopenia** | 2(6.3%) | 4(7.8%) | 1(2.7%) | 2(6.3%) | 0.40 |
| **Anemia** | 5(15.6%) | 10(19.6%) | 1(2.7%) | 2(6.3%) | 0.28 |

**Fig 1.**
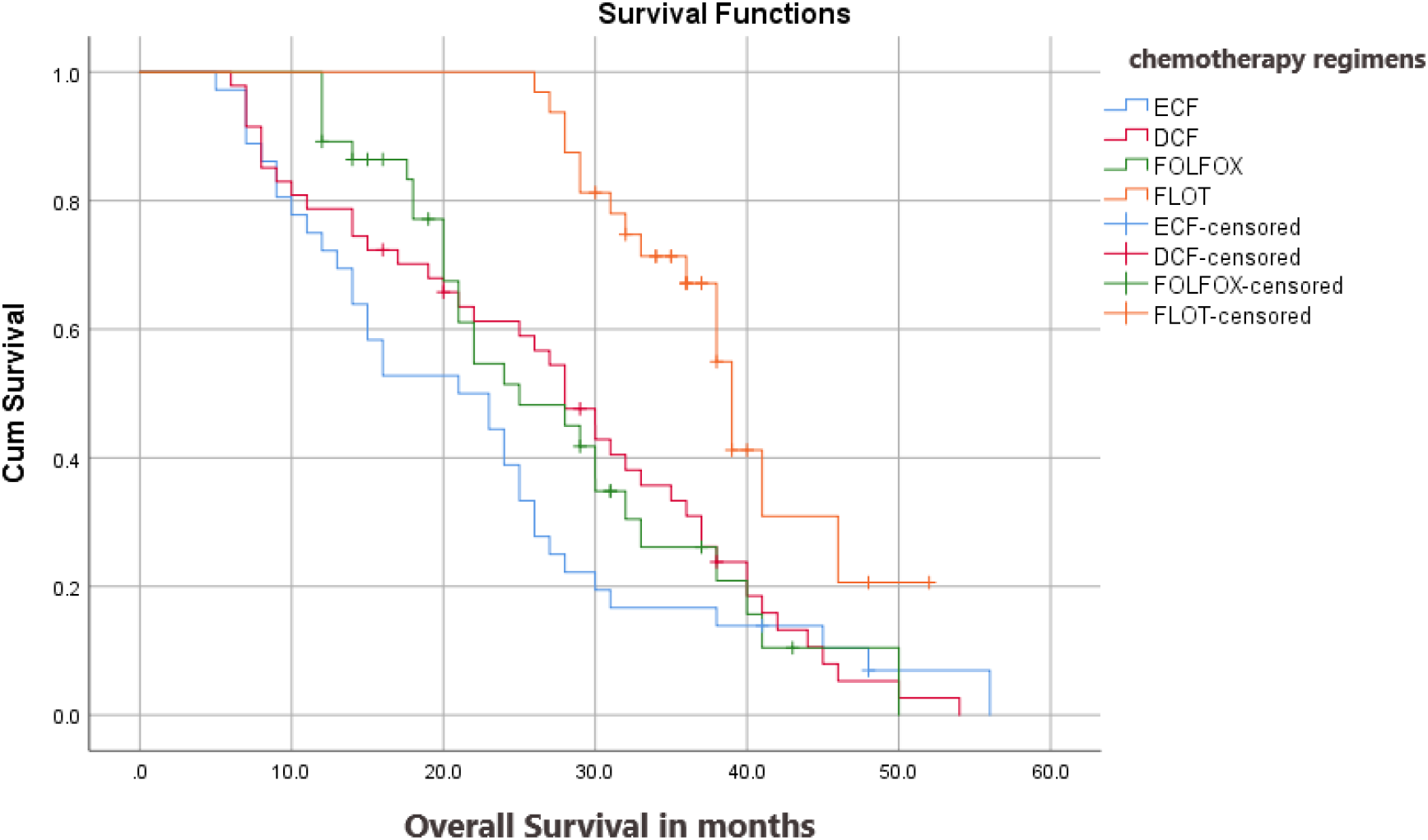
Kaplan-Meier plot of overall survival.

**Fig 2.**
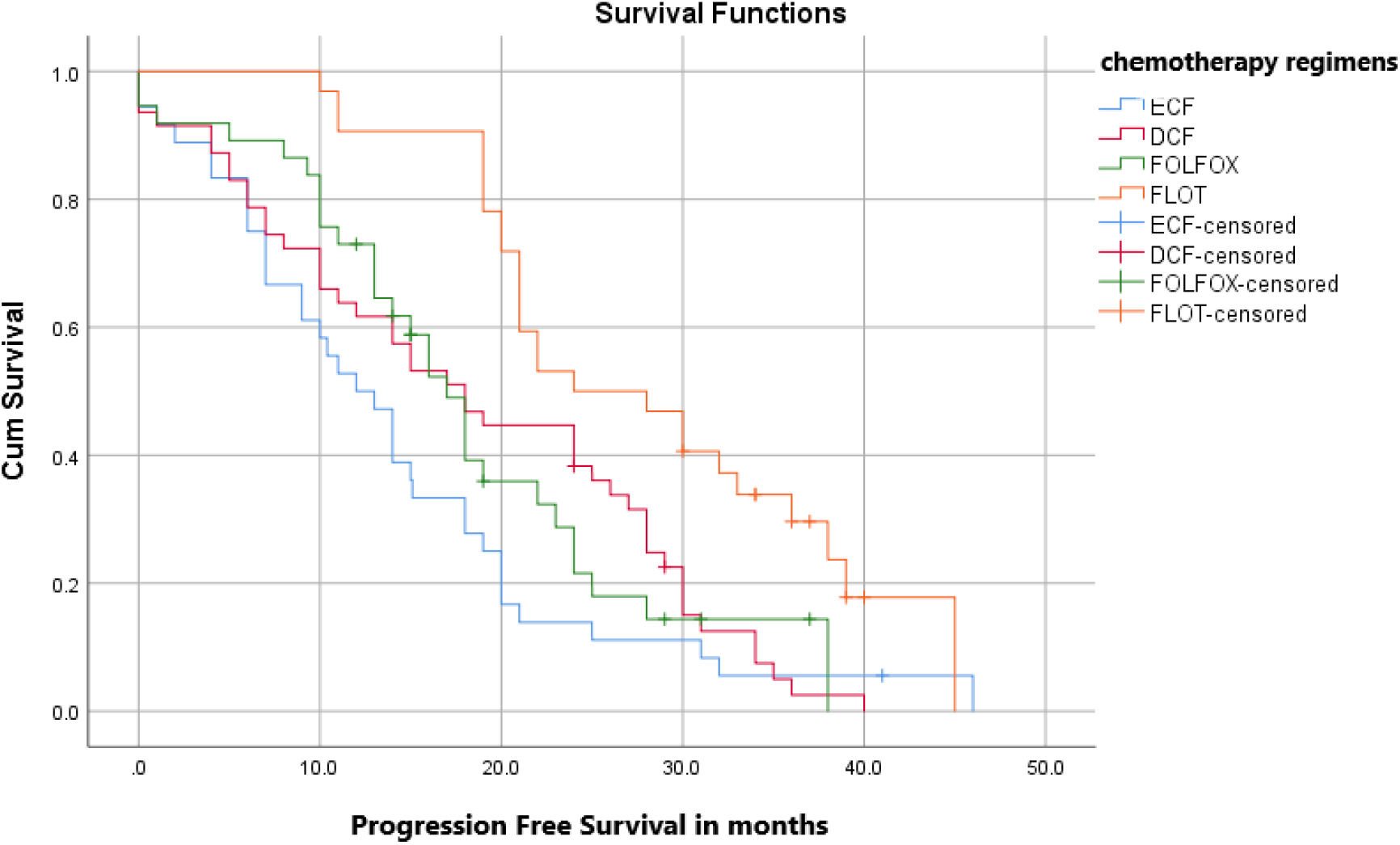
Kaplan-Meier plot of progression-free survival.

### Toxicity

All 3-4 grades of hematological and non-hematological toxicities were reported in Table 5. Grade 3-4 neutropenia was noted in 11(34.3%), 26(51%),6(16.2%), 12(37.2%) of patients receiving ECF, DCF, FOLFOX and FLOT regimens, respectively. The most-reported 3-4 grade of non-hematological toxicities were occurrences of gastrointestinal adverse effects such as nausea and diarrhea noted in 30(19.7%) and 26(17.1%) of the enrolled patients. DCF regimen represented a higher rate of toxicity in comparison with other chemotherapy regimens.

### Multivariate analyses

The results of univariate analysis by Cox regression revealed that several factors including ECOG performance status, family history of cancer, disease histopathology, cancer stage, tumor size, having other organs’ metastasis, site of metastasis, resection rate, and response to chemotherapy had a significant correlation with both PFS and OS. (Data were summarized in supplement Table 1)

As shown in supplement Table 1, the results of multivariate analysis by Cox regression showed a significant correlation between diminishing in OS by variants such as having poorly-differentiated pathology (HR=2.14, p-value=0.002), the lack of response to chemotherapy (defined as progressive disease or stable disease) (HR=63.02, p-value<0.001) and R1 resection (HR=12.63, p-value<0.001).

On the other hand, several patients’ factors such as being underweight (BMI<18) (HR=2.06, p-value=0.02), having ECOG performance status >1 (HR=3.57, p-value=0.02), having poorly-differentiated pathology (HR=1.7, p-value=0.02), no response to chemotherapy (define as a progression disease or stable disease) (HR=225.17, p-value<0.001) and R1 resection (HR=3.34, p-vaue <0.001) had lower chance of PFS.

**Supplement Table 1.**
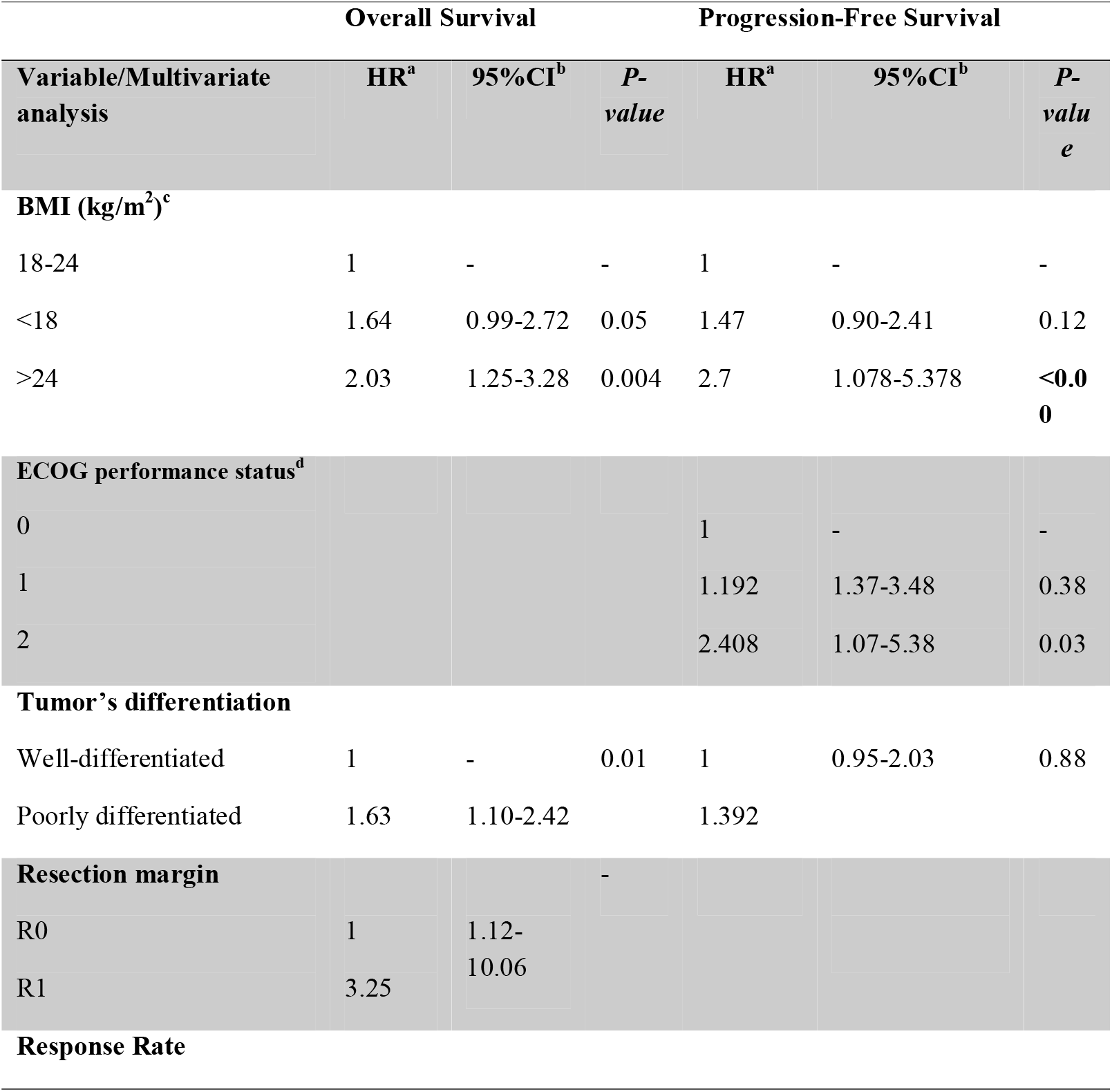
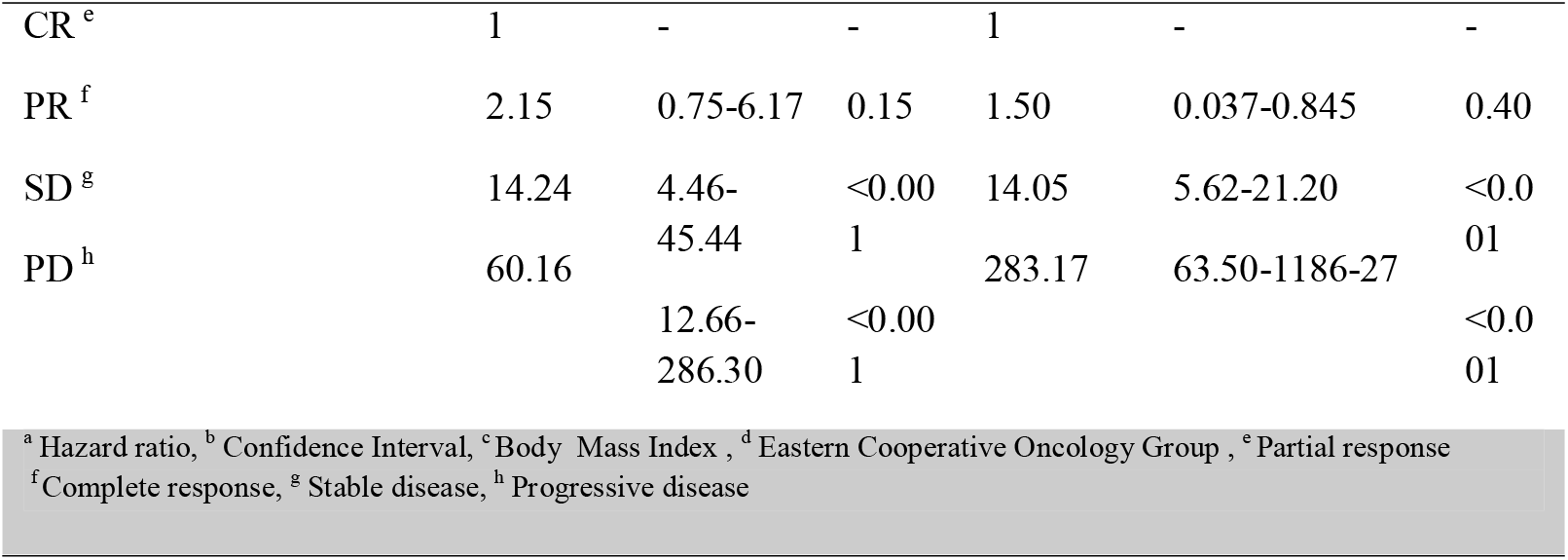
Multivariate analysis of overall survival and progression-free survival.

## Discussion

In this retrospective study, for the first time in the Iranian population, we evaluated the most common administrative chemotherapy regimens for the perioperative protocol of management for gastric cancer patients with resectable tumors, including ECF, DCF, FOLFOX, and FLOT. Based on our implication, the FLOT regimen ascertained significant improvement in patients’ OS and PFS in comparison with the other regimens.

The predominance efficiency of perioperative FLOT regimen in managing resectable gastric cancer has been demonstrated in the FLOT-AIO4 study which had been compared FLOT and ECF perioperative chemotherapy regimens in 356 and 360 patients with resectable gastric cancer, respectively[14]. In this study, the FLOT regimen was superior in demonstrating either OS up to 50 months or PFS up to 32 months than the ECF regimen[14]. Additionally, *Schulz et al*. phase 2 trial with 50 patients who had received the perioperative FLOT regimen showed that the FLOT regimen improved PFS (32.9 months) significantly in comparison with the surgery alone[22]. Similarly, our study results revealed that the FLOT regimen had superiority in increasing both PFS and OS in comparison with a reference regimen (ECF regimen). However, the median Overall and progression survival time in our study were 39 and 24 months, respectively. Which were lower than the previous studies[14.22], ^1^possibly due to the effect of sample size. Moreover, according to *Malekzadeh et al*. study, most of the Iranian patients suffering from gastric cancer were diagnosed in the advanced level of tumors that can be directly attributed to poor survival time[23]. The most two noted reasons for delayed recognition of Iranian gastric cancer patients were patients’ refusal and economic problems[23].

It is worth mentioning that, as opposed to other noted median PFS time for FLOT chemotherapy regimen[13,14,22], the one-year and two-year reported OS rate, as well as indicators such as ORR and DCR in FLOT regimen, was higher in our study than other similar studies[13,14,22]. ^1^In the present study, comparing the FLOT regimen with other chemotherapy regimens has shown the superiority of this regimen not only by improving OS and PFS period but also by enhancing survival rate and R0 resection rate. R0 resection rate was considered as an endpoint in our investigation and demonstrated to be as high as a 93.8% resection rate for the FLOT regimen. This resection rate for the FLOT regimen was similar to *Wang K*. *et al* retrospective study with a 91% resection rate in 40 included patients[13], and higher than the resection rate reported by *Schulz C et al*. study which was 86% in 50 patients[22].

When it comes to the DCF regimen, the median OS and PFS were reported 28 months and 18months, respectively. The results were compatible with similar studies that were conducted to evaluate the efficiency of the perioperative DCF regimen[25,25]. Nevertheless, in the retrospective survey conducted by *Fiteni et al*.[26], the reported median OS (57 months) and three-year survival rate (60%) were higher than our results or even other similar studies[24,25] which can be due to longer follow-up duration (near 10 years) and more enrolled patients (62 patients) in comparison to our study.

In the resemblance with recently published meta-analysis[27,28], our cox-regression analysis demonstrated that there was a significant difference between OS enhancement by ECF and DCF regimens. The superiority of taxane-based regimen over non-taxane-containing chemotherapy was confirmed by *Chaudhuri et al*. not only inoperable gastric cancer patients but also in the metastatic situation[29]. Besides, we found that the PFS of patients receiving the DCF regimen was higher than the ECF regimen, despite non-statistically significant results. Even though, the taxane-based regimen has shown improvement in PFS rather than an anthracycline-based regimen; however, the difference was not statically significant[28]. To put all data into a nutshell, despite several data over the increasing PFS by both DCF and ECF regimens[6,26,29], DCF regimen remained the superior regimen in the perioperative setting of gastric cancer’s management due to enhancement in other survival indices such as median time of overall, progression-free survival and overall response rate.

We found that in the second place after the FLOT regimen, the DCF regimen led to an increase in median survival time, DCR, ORR, and R0 resection rate in comparison with FOLFOX and ECF regimens; however, the maximum level of toxicity has been detected by DCF regimen. Despite the similar resection rate reported between DCF and FLOT regimens, by considering other survival endpoints such as median survival time, ORR, and toxicity, the DCF regimen seems to be inferior in comparison with the FLOT chemotherapy regimen. Furthermore, introducing the DCF regimen as an optimal regimen is not reasonable.

Concerning the FOLFOX regimen, the numbers of our survival analysis results were consistent with other studies that utilized a variety of types of FOLFOX regimen as perioperative chemotherapy regimens [10,12,30,31]. To illustrate this resemblance, the retrospective cohort study conducted *by Sun et al*. FOLFOX4 regimen showed 29-month OS time liken to our noted 25-month median OS in our study[10].

Moreover, our reported one and two-year OS, as well as ORR (75.5%) and DCR (89.1%), was comparable with other similar studies administrated FOLFOX7 and modified-FOLFOX6 regimens[12,30]. However, our21% OS rate after three-year follow-up was in disagreement with the approximate amount of 60% which was reported in the previous studies[10,12,29,30]. The lower OS rate in patients who had received FOLFOX regimen in our study can be attributed to the higher median age of our enrolled patients (60 years old) in comparison with the other important studies with less than 50 years old patients recruitment[10,12].

Regarding the efficacy of the ECF regimen, the results of one-year survival in our study (71.9%) were in accordance with both studies including FLOT-AIO4 and MAGIC that were successful in showing the almost 60-70% one-year survival rate during their follow-up[6,14]. In a retrospective cohort study conducted by *Achili et al*. ECF as a perioperative chemotherapy regimen, the ORR and DCR rates were reported 37% and 42% accordingly, which is less than our study with an ORR rate of 56% and DCR rate of 87.6%[32]. However, in our study, 21 months median OS and 14 months PFS were lower than the aforementioned study which had 25.7 months PFS and 36.6 months OS[32]. This controversy is explainable by less number of enrolled patients and a shorter period of follow-up in our study. In addition, Iranian patients were diagnosed at a higher stage of the disease which may lead to a lower chance of survival[12,23,30].

In consensus with the aforementioned results of our study about the inferiority of the ECF regime, this regimen showed the lowest R0 resection rate. This R0 resection rate was 75% which was comparable to other reported Ro resection rates for the ECF regimen[6,14]. By considering all factors related to survival including median PFS, OS, ORR, and R0 resection rate, the ECF regimen failed to represent the optimum efficiency in comparison to other regimens.

Regarding the toxicity of chemotherapy regimens as a secondary endpoint in the study, we could show that the DCF regimen was significantly associated with more grade 3-4 toxicities such as neutropenia and mucositis in comparison with another regimen. The reported grade 3-4 hematological toxicity especially neutropenia in our study were in accordance with other reported DCF regimen’s toxicity rate which was around 31-87%[24,26,34,35]. Besides, the rate of 3-4 grade mucositis and hospitalization for the DCF regimen were 21% and 52% respectively, similar to other published reports[34-36].

On the other hand, the grade 3-4 neutropenia incidence rate for the FLOT regimen was reported between 30-51%[14,22,37], in a range revealed by our findings. Although the rate of 3-4 grade mucositis (16%) was higher than other similar studies[14,22] due to possible differences in inappropriate oral hygiene in Iranian patients[38], as an important risk factor of mucositis[39]. The hospitalization rate for our study was 37% as opposed to the FLOT-ATIO4 trial with a 25% rate.[14] This difference can arise by inappropriate use of G-CSF by some physicians to the prevention of chemotherapy toxicity in our institute[40].

Like the other similar reports[60,31,41], the FOLFOX regimen had the lowest rate of toxicities among the other three regimens by inducing 16% grade 3-4 neutropenia and 8.5% grade 3-4 mucositis.

It is worth mentioning that another advantage of using oxaliplatin-based chemotherapy regimens like FLOT and FOLFOX is a lower rate of adverse effects rather than a cisplatin-based regimen like DCF and ECF. In addition, oxaliplatin-based regimens maintain the same or even better effectiveness overall. This issue is well demonstrated in the Meta-analysis which was conducted by *Huang J et al*.to compare the effectiveness and safety of oxaliplatin-based and cisplatin-based therapy in advanced gastric cancer[42] and demonstrated that there is no difference in terms of the effectiveness of factors such as OS, PFS, and ORR between the oxaliplatin and cisplatin-based chemotherapy[41]. However, the oxaliplatin-based regimen was associated with less adverse toxicity except for neurotoxicity.

In multivariate regression analysis, the histopathological tumor type showed a significant correlation with patients’ survival time (both OS and PFS) in a way that patients with the well-differentiated subtype had improved OS and PFS remarkably. These finding has been noted in other similar studies[43], in which the histopathology of the gastric tumor was introduced as an independent risk factor of gastric cancer patients’ OS. Also, in agreement with our findings, *Castellanos et al*. retrospective study[24], and *a Li et al*. meta-analysis (2018) reached the conclusion that the response to chemotherapy and achieving to R0 resection rate significantly increased the time of survival in gastric cancer patients[44].

Body mass index was detected as a prognostic factor for PFS and OS according to multivariate and univariate analysis as well. Along with, underweight patients and overweight patients considerably had a lower chance of survival in comparison with normal BMI patients suffering from gastric cancer. In a results analysis of a large cohort study that recruited 7700 gastric cancer patients, underweight patients showed a lower duration of survival, although obese patients had a better survival[45]. It is worth mentioning that malnutrition and less reserve of adipose tissues were repeatedly noted as the prognostic factors of poor survival in cancer patients and could diminish the anti-tumor activity of the immune system[45,46]. Furthermore, it will be recommended that nutritional support before and during chemotherapy should be considered for the underweight patient suffering from gastric cancer. In addition, some studies suggested that lower BMI was associated with a higher rate of lymph node metastasis and a more advanced tumor stage that is considered as a poor prognosis factor in gastric cancer[46,47].

To our knowledge, this is one of the first studies performed to compare the efficacy and tolerability of most common perioperative chemotherapy regimens in the managing of resectable gastric cancer in the Iranian population. *Mousavi et al*. in 2015 evaluated the efficacy of several most prescribed chemotherapy regimens in resectable and metastatic gastric cancer patients lived in the north of Iran and concluded that the DCF regimen prolonged patients’ survival time more than ECF, DOX, and Xeloda -based chemotherapy[48]. Due to a high prevalence of gastric cancer in the countries such as Iran and a lack of definite national and international guidelines to recommend the best therapeutic options[23,48], the necessity of conducting a comparative study among chemotherapeutic regimens is crucial.

Our study suffered from several limitations. The First FLOT regimen has recently been used in our country and for completing the ultimate documents, more follow-up duration and more patients who will be treated by FLOT regimen is necessary. Second, due to the retrospective methodology of the study, meticulous documentation and data gathering were challenging. To overcome the limitations and verify our results, a multicenter randomized clinical trial in the Iranian population for collecting comprehensive and accurate data to decide on the optimal first-line chemotherapy regimen for the treatment of resectable gastric cancer as perioperative modalities is recommended.

## Conclusion

We demonstrated that the FLOT chemotherapy regimen significantly enhanced survival parameters including OS and PFS and additionally exerted a higher rate of R0 resection and overall response with a tolerable toxicity profile in comparison with other included chemotherapy regimens. Conducting large and well-designed clinical trials to confirm the assertion is crucial.

## Data Availability

All data are available upon request.

## Acknowledgments

We acknowledge by Isfahan University of Medical Sciences for funding this research and all health care professions in Omid hospital helped us in data gathering and patients’ follow-up procedures.

## Declarations

### Ethics approval

The study was approved by the Ethics Committee of Isfahan University of Medical Sciences with the Iranian approval ID of IR.MUI.RESEARCH.REC.1398.167.

### Availability of data and material

available upon request.

### Code availability

available upon request.

### Contributor ship

PF: Data gathering, data analysis and writing of the article, AZ: Design of the study, data analyses, manuscript preparation, and editing. AS and MS: Data gathering and study design. PD and RR: Data processing and manuscript editing. The final version of the article was approved by all authors before publication.

